# Context-dependent facial-expression patterns during affective film viewing in patients with bipolar depression

**DOI:** 10.64898/2026.07.19.26358451

**Authors:** Eunhwi Lee, Sung Hun Sim, Chanhee Park, Hyuntae Kim, Woo-Young Ahn, C. Hyung Keun Park

## Abstract

**Background:** Emotion dysregulation is a core feature of bipolar disorder (BD), yet its behavioral expression during depressive episodes, and potential differences between its types, BD-I and BD-II, remain unclear. This study used automated facial-expression analysis during naturalistic affective film viewing to examine subtype-specific and context-dependent emotional responding in bipolar depression.

**Methods:** The sample included 135 participants: 69 healthy controls and 66 patients with BD (BD-I, 23; BD-II, 43). Participants viewed nine emotionally evocative film clips spanning negative, positive, neutral, and socially threatening contexts, while their facial expressions were continuously recorded and quantified using computer vision-based facial-expression analysis.

**Results:** Patients with BD-I showed a distinct, context-dependent facial-expression profile, characterized by greater negative responses across multiple contexts than other groups. Specifically, they showed increased sadness during sad, reward, and amusing clips, and elevated anger during sad and neutral clips. In socially threatening contexts, BD-I participants showed a multivalent pattern of elevated anger, fear, and joy, suggesting poorly coordinated or context-incongruent affective expression. In contrast, BD-II participants did not differ significantly from healthy controls on any emotion, despite depressive symptom severity comparable to BD-I participants.

**Conclusions:** These findings suggest that facial-expression patterns in bipolar depression differ across subtypes. BD-I may be characterized by heightened negative reactivity and altered context-appropriate modulation of emotional expression, whereas BD-II may not show comparable alterations in overt facial output. Automated facial-expression analysis during naturalistic stimulation may provide a useful behavioral marker for characterizing subtype-specific affective disturbance in bipolar depression and related psychopathology.

## 1. Introduction

Bipolar disorder (BD) is a recurrent and highly impairing mood disorder characterized by alterations in emotional responding across mood states (Townsend & Altshuler, 2012). These alterations may affect daily functioning by disrupting emotion regulation, inter-personal communication, and social engagement. Such disturbances are associated with poor functional outcomes, including lower quality of life and psychosocial functioning in remitted bipolar I disorder (BD-I) (Johnson et al., 2016). BD is also associated with markedly elevated risk of suicide across the illness course (Dome et al., 2019).

Despite their clinical relevance, disturbances in emotional responses in BD have not been captured consistently at the behavioral level. Conventional self-report or symptom-based measures may not be sufficiently sensitive to detect subtle or context-dependent abnormalities of emotional responses in BD, as suggested by studies of patients with BD in full or partial remission (Broch-Due et al., 2018; Kjærstad et al., 2020). Accordingly, there is a need for more sensitive and objective approaches that can characterize these responses.

One potentially useful approach is the assessment of spontaneous facial expression during emotional stimulation. Facial expressions provide an observable behavioral index of emotional responses, and automated facial-expression analysis allows these responses to be quantified continuously and objectively during dynamic stimuli. Compared with manual facial coding, which is labor-intensive and susceptible to variability across raters, computer vision-based approaches can provide standardized estimates of facial action units and emotion-related expressions over time. These approaches have been increasingly well-validated for facial-expression recognition and continuous affect estimation (Bartlett et al., 1999; Haines et al., 2019; Stöckli et al., 2018), with applications across clinical research on pain (Sikka et al., 2015), depression (Haque et al., 2018; Mao et al., 2023), and other psychiatric symptoms (Bilgrami et al., 2025; Gupta et al., 2019; Martin et al., 2024).

Previous studies suggest that objective behavioral indices such as facial expressions may detect subtle abnormalities in emotion processing in BD more sensitively than self-report measures alone, including cases in which group differences are not clearly evident in subjective emotional ratings (Broch-Due et al., 2018; Kjærstad et al., 2020). In BD, studies using eye-tracking or facial-expression analysis have reported altered gaze patterns and subtle differences in facial-expression responses during exposure to emotional pictures or film clips (Broch-Due et al., 2018; Kjærstad et al., 2020). For example, these patients show atypical facial patterns, including stronger expressions to neutral pictures but reduced expressions to unpleasant pictures, as well as increased fearful facial expressions across emotional film clips in remitted samples (Broch-Due et al., 2018; Kjærstad et al., 2020). Other studies have also reported deficits in related aspects of emotion processing, such as facial emotion recognition in BD (Derntl et al., 2009; Martino et al., 2011). Together, these findings support the value of using facial expression markers to characterize emotion-related dysfunction in BD. However, several questions remain unresolved regarding how such abnormalities vary across bipolar subtypes and how facial expressions can be used to objectively evaluate emotion-related dysfunctions in naturalistic emotional contexts.

A particularly important issue is that bipolar I and bipolar II disorders (BD-II) should not be assumed to show identical emotional profiles, as the two subtypes differ in clinical course and depressive burden (Brancati et al., 2023; Judd et al., 2003). Compared with BD-I, BD-II is more often characterized by depressive onset, recurrent depressive episodes, depressive-predominant polarity, and a greater proportion of illness time spent in depressive states (Brancati et al., 2023; Judd et al., 2003). These distinctions suggest that emotional responding during bipolar depression may not be uniform across bipolar subtypes. Therefore, this warrants direct comparison rather than treating BD as a homogeneous population. Nevertheless, most prior studies have either pooled these patients into a single diagnostic group, or have not directly compared BD-I and BD-II, leaving it unclear whether the two subtypes show distinguishable patterns of spontaneous emotional responding (Broch-Due et al., 2018; Kjærstad et al., 2020). Even when subtype differences have been considered, such studies have typically been conducted in euthymic or otherwise symptomatically stable samples, rather than in patients who are experiencing a current major depressive episode (Samame et al., 2012). This is a particularly important limitation because major depressive episodes represent a clinically central and burdensome phase of BD, given the predominance of depressive symptoms in the longitudinal course of the illness (Brancati et al., 2023; Judd et al., 2003). Therefore, subtype-related differences in emotional responding may be especially relevant at that stage.

Furthermore, prior work has only provided limited insight into how emotional responses are expressed in more naturalistic settings. Much of the literature relevant to emotion processing in BD has relied on static pictures, emotion-recognition tasks, or broader social-cognitive paradigms rather than directly examining emotion responsivity during dynamic emotional contexts (Broch-Due et al., 2018; Derntl et al., 2009; Martino et al., 2011; Samame et al., 2012). Although Kjærstad et al. (2020) examined eye gaze and facial displays during emotional film clips, their study focused on remitted patients with BD rather than patients experiencing a current depressive episode. Thus, it remains unclear how emotion is spontaneously expressed in response to more naturalistic, temporally unfolding stimuli during bipolar depression. This limitation is particularly relevant when the goal is to characterize emotion responding patterns across diverse contexts that more closely resemble real-world affective experiences.

To address these gaps, the present study examined facial expressions during emotional film clips in patients with BD-I or BD-II, all of whom were experiencing a current major depressive episode, and healthy controls (HCs) using automated facial coding. This design enabled the testing of subtype-related differences in emotional expression under clinically relevant and more naturalistic conditions than the static or constrained paradigms. By using emotionally evocative film clips and automated facial coding, the present study was able to assess how facial responses unfolded across different emotional contexts. It was expected that BD-I and BD-II would show different patterns of facial expression across emotional contexts, but given the limited evidence base, the specific nature of those differences was treated as an open empirical question.

## 2. Methods

### 2.1. Participants

Participants were recruited from March 2024 to December 2024 as part of a broader ongoing project on emotional memory and attentional bias in BD. Patients were recruited from the Department of Psychiatry at Asan Medical Center, and HCs were recruited through community advertisements. Inclusion criteria were: age between 18 and 64 years, at least a high school education, and, for the patient group, a diagnosis of BD-I or BD-II with a current major depressive episode confirmed by the Mini-International Neuropsychiatric Interview (MINI; Sheehan et al., 1998). Exclusion criteria were: substance abuse or dependence within the past six months, intellectual disability, lifetime attention-deficit/hyperactivity disorder, head trauma involving loss of consciousness for more than three minutes, uncontrolled neurological illness, unstable medical illness affecting brain function, inability to understand Korean, color-vision deficiency, and strabismus. The study was approved by the Institutional Review Board of Asan Medical Center (IRB No. 2023-0178), and written informed consent was obtained from all participants. Of the 140 participants recruited, five were excluded from the analysis: two due to an insufficient number of valid columns to calculate the ratio of emotional evidence, one because the participant was later found not to meet the patient eligibility criteria, and two because the recordings were not saved due to experimenter error. The final sample consisted of 135 participants (69 HCs, 23 BD-I, and 43 BD-II).

### 2.2. Measures

Depressive symptoms were assessed using the Patient Health Questionnaire-9 (PHQ-9; Kroenke et al., 2001), and anxiety symptoms were measured with the Generalized Anxiety Disorder-7 scale (GAD-7; Spitzer et al., 2006). Suicidal ideation was evaluated using the Columbia-Suicide Severity Rating Scale (C-SSRS; Posner et al., 2011). Detailed descriptions of the measures are provided in **Supplementary Material 1**.

### 2.3. Experimental paradigm

The experimental paradigm was adapted from the emotional film-clip task used by Kjærstad et al. (2020), with clips newly selected from Korean-language materials to improve cultural and contextual relevance for Korean participants. A total of nine film clips were used (see **Table 1** for a description of each film clip). Compared with the original paradigm, anger- and amusement-eliciting clips were additionally included to broaden the range of emotional contexts represented in the task. For each target emotional category, two candidate clips were evaluated by an independent sample of 31 individuals who did not participate in the main experiment. The clip showing better agreement with the intended emotion and a higher rating on that emotion was selected as the final stimulus. Detailed information on the stimulus-selection procedure is provided in **Supplementary Material 2**.

**Table 1.** Final stimulus set and brief narratives of the emotional film clips Film clip Narrative.

| Film clip | Narrative |
| --- | --- |
| Amusing | A comedian performs physical comedy on stage. |
| Anger | A woman recounts a painful personal experience from a historical period of oppression. |
| Happy | A baby smiles brightly and repeatedly. |
| Neutral | A person pours tea into a cup. |
| Sad | Family members who were separated in childhood unexpectedly recognize and reunite with each other. |
| Racing | A car chase unfolds at high speed. |
| Risk taking | A person climbs along the exterior of a building to escape from a fire. |
| Socially<br>anxious | A person struggles to suppress the need to use the bathroom during an exam. |
| Winning | A short-track speed skater makes a dramatic comeback and wins the final race at the Winter Olympics. |

Each emotional film clip was presented for 90 s in a randomized order across participants. Stimulus presentation was controlled using PsychoPy (v2022.2.3), an open-source software package for behavioral experiments (Peirce et al., 2019). To minimize carry-over effects from the preceding clip, a 30-s visually neutral wave clip was inserted between film clips, consistent with prior emotion-induction paradigms that used neutral or washout intervals for this purpose (Davies et al., 2013; Leppanen et al., 2017). Accordingly, the total duration of the task was 17 min 30 s. Participants’ facial expressions were recorded throughout the task using a Logitech BRIO 4K Pro webcam.

Facial expressions were analyzed using automated facial expression analysis (FEA) implemented in iMotions (version 10.0; iMotions A/S, Copenhagen, Denmark), which incorporates the Emotient FACET engine, a commercialized version of the Computer Expression Recognition Toolbox (CERT), which has been validated in previous work for automated facial detection and expression recognition, supporting their use for frame-by-frame quantification of facial responses (Littlewort et al., 2011; Stöckli et al., 2018). This computer vision tool applies facial action unit (AU) detection algorithms based on the Facial Action Coding System (FACS; Ekman & Friesen, 1978) to quantify facial muscle movements on a frame-by-frame basis. AU patterns are then used to estimate the likelihood of discrete affective states, including joy, anger, surprise, fear, contempt, disgust, sadness, and neutral expression, as well as composite indices of positive and negative affect. These outputs are expressed as continuous probability scores ranging from 0 to 1, reflecting the likelihood that a given emotion is present in each frame.

Prior to the film clip task, participants viewed a seven second fixation cross to establish an individual resting baseline. For each participant, the median probability score across all fixation frames was computed and subsequently subtracted from the corresponding frame-level scores recorded throughout the task, following the iMotions guideline (iMotions, 2026). This procedure corrects for individual differences in resting facial muscle tone and webcam-related artifacts.

For each emotion within each clip, the primary dependent variable was defined as the proportion of frames in which the baseline-corrected facial expression probability score equaled or exceeded 0.5, as recommended by iMotions for moderate changes in facial expressions (iMotions, 2026). This measure reflects the proportion of time during which a given emotion was detectably expressed above the individual baseline. This approach is consistent with prior work using affective computing and facial expression analysis, where threshold-based percentile metrics are used to capture the occurrence and duration of emotional expressions (Korn & Rees, 2019).

### 2.4. Statistical analysis

Group differences in facial expression were examined using one-way analysis of variance (ANOVA) with three between-subjects groups (HCs, BD-II, BD-I). Analysis was conducted at two levels: across all nine emotionally evocative film clips combined, using each participant’s mean value across clips, and separately for each individual clip. Significant omnibus effects were followed up with Tukey’s honestly significant differences (HSD) post-hoc comparisons. Effect sizes are reported as partial eta-squared (η²p). The significance threshold was set at α = .05.

As a supplementary analysis, to account for the repeated-measures structure of the clip-level data, and to assess whether the ANOVA findings were robust when all clips were modeled simultaneously, linear mixed-effects models (LME) were estimated for each emotion channel with group, video, and their interaction entered as fixed effects, and a participant included as a random intercept. An additional model including demographic and clinical variables as co-variates was estimated to assess the robustness of the results. All analysis was conducted in R (version 4.3.3).

## 3. Results

### 3.1. Comparison of participant characteristics across BD-I, BD-II, and HCs

Demographic and clinical characteristics of the participants are summarized in **Table 2**. The three groups did not differ significantly in age or sex distribution. HCs had more years of education than both patient groups, and familial history of psychiatric disorders was more frequent in BD-I and BD-II than in HCs. As expected, both patient groups showed higher depressive and anxiety symptoms than HCs. Importantly, depressive symptom severity was comparable between BD-I and BD-II, indicating that subsequent subtype-related differences in facial expression were unlikely to be explained by differences in overall depressive status. C-SSRS severity was highest in BD-I, followed by BD-II and HCs.

**Table 2.** Differences among healthy controls, patients with bipolar II and bipolar I depression.

| Variables | HC (N = 69) | BD-II (N = 43) | BD-I (N = 23) | $F/\chi^2$ | $p$ |
| --- | --- | --- | --- | --- | --- |
| Age, years (SD) | 32.72 (7.05) a | 31.26 (10.42) a | 35.00 (12.50) a | 1.22 | 0.30 |
| Female, n (%) | 52 (75.4) a | 32 (74.4) a | 18 (78.3) a | 0.12 | 0.94 |
| Education, years (SD) | 16.26 (1.49) a | 14.88 (2.51) b | 14.70 (1.82) b | 9.56 | <0.001 |
| Familial history of psychiatric disorders, n (%) | 10 (14.5) a | 18 (41.9) b | 11 (47.8) b | 14.50 | <0.001 |
| **Psychiatric measures** |  |  |  |  |  |
| PHQ-9 (SD) | 1.84 (2.40) a | 20.05 (6.16) b | 21.39 (4.72) b | 314.23 | <0.001 |
| GAD-7 (SD) | 0.90 (1.62) a | 14.47 (5.19) b | 15.70 (4.18) b | 258.03 | <0.001 |
| C-SSRS Severity (SD) | 0.00 (0.00) a | 2.30 (1.52) b | 3.17 (1.77) c | 95.40 | <0.001 |

\* HC, healthy control; BD-II, bipolar II depression; BD-I, bipolar I depression; SD, standard deviation; PHQ-9, Patient Health Questionnaire-9; GAD-7, Generalized Anxiety Disorder-7; C-SSRS, Columbia-Suicide Severity Rating Scale. Different subscripts within rows indicate statistically significant differences at $p < 0.05$ .

### 3.2. Associations between facial expression and clinical symptom severity

Before examining group differences in facial expression, associations between facial expression proportions and demographic and clinical variables were tested using Spearman correlations for age, education, C-SSRS severity, PHQ-9, and GAD-7; and Mann–Whitney U tests for sex and family history. No associations remained significant after false discovery rate (FDR) correction. The same null pattern was observed in within-group analysis conducted separately for each group. These findings suggest that the group differences in facial expression proportions reported below were not systematically explained by concurrent variation in demographic or clinical characteristics.

### 3.3. Group differences in facial expression

In terms of overall emotion expression across the nine film clips combined, a significant group difference was observed for sadness, with BD-I participants showing sadness responses more frequently than HCs (F(2, 132) = 3.61, p = 0.030, η²p = 0.052; BD-I: M = 0.310, SD = 0.254; HCs: M = 0.184, SD = 0.185; Tukey HSD: p = 0.026). No other emotion channel reached significance at the overall level; anger showed a non-significant trend, with BD-I participants displaying anger more frequently than HCs (BD-I: M = 0.400; Control: M = 0.256; p = 0.091).

At the individual clip level, the most consistent pattern involved elevated sadness in BD-I across multiple emotional contexts. During a clip depicting a sad scenario (**Figure 1A**), BD-I participants showed sadness responses significantly more frequently than both controls and BD-II participants (F(2, 132) = 7.22, p = 0.001, η²p = 0.099; BD-I: M = 0.486, SD = 0.362; BD-II: M = 0.241, SD = 0.299; Control: M = 0.225, SD = 0.263; Tukey HSD: BD-I > Control, p = 0.001; BD-I > BD-II, p = 0.004). This pattern extended to a reward-related context: during a clip depicting a winning scenario (**Figure 2A**), BD-I participants again showed more frequent sadness responses than both controls and BD-II participants (F(2, 132) = 5.27, p = 0.006, η²p = 0.074; BD-I: M = 0.291, SD = 0.309; BD-II: M = 0.155, SD = 0.228; HCs: M = 0.119, SD = 0.178; Tukey HSD: BD-I > HCs, p = 0.004; BD-I > BD-II, p = 0.049). Similarly, during an amusing clip (**Figure 2B**), BD-I participants showed sadness responses more frequently than HCs (F(2, 132) = 3.23, p = 0.043, η²p = 0.047; BD-I: M = 0.221, SD = 0.271; HCs: M = 0.089, SD = 0.183; p = 0.050).

**Figure 1.**
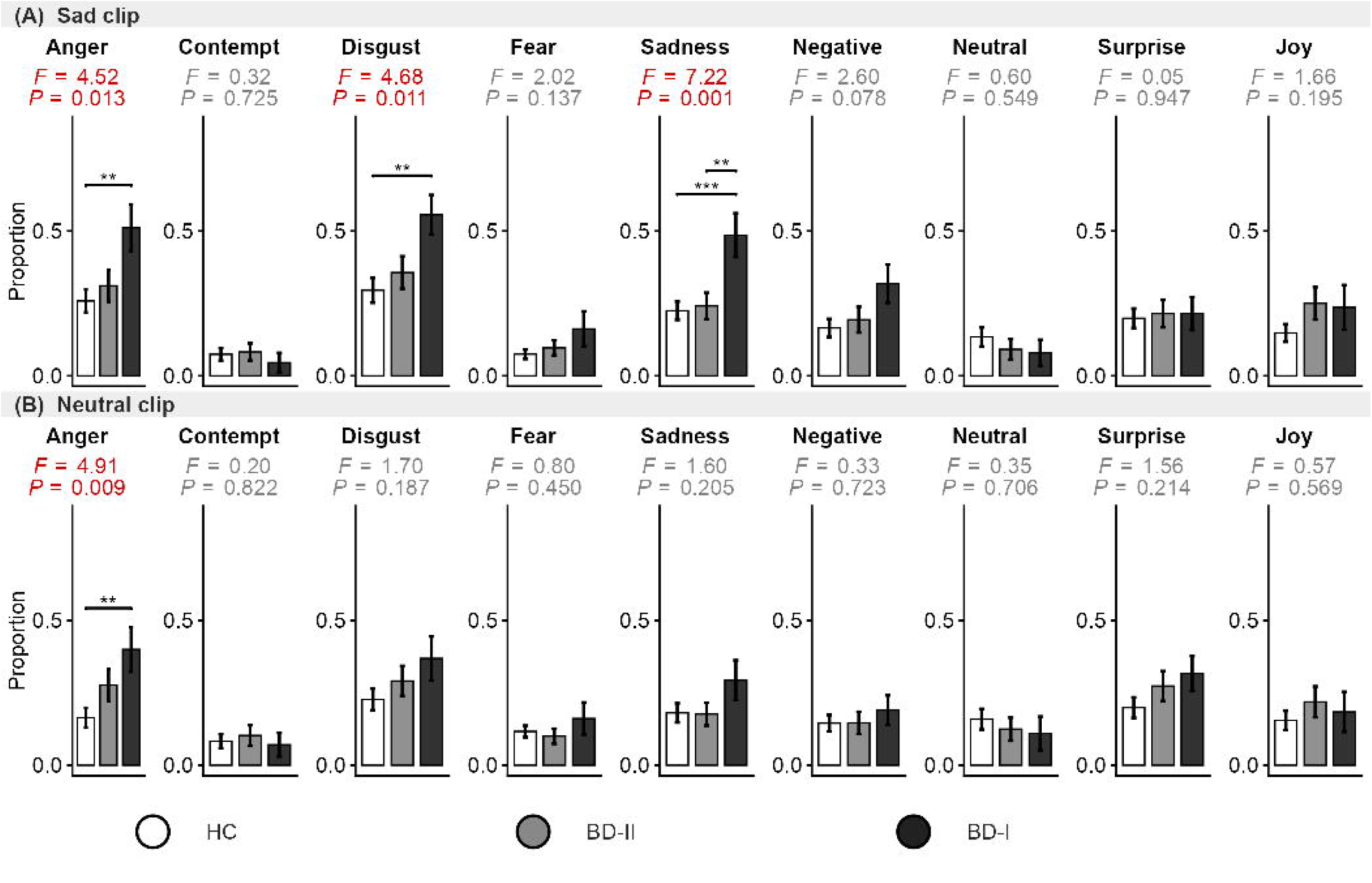
Facial expression proportions during sad and neutral film clips. Bar graphs show the mean values of the groups with standard error bars for each emotion channel during (A) a sad film clip, and (B) a neutral film clip. The dependent variable is the proportion of frames in which the baseline-corrected facial expression probability score equaled or exceeded 0.5. Significant between-group differences (one-way ANOVA, p < .05) are indicated by red F and p values; significant post-hoc pairwise comparisons (Tukey HSD) are denoted by brackets (* p < .05, ** p < .01, *** p < .001). BD-I, bipolar disorder type I; BD-II, bipolar disorder type II.

**Figure 2.**
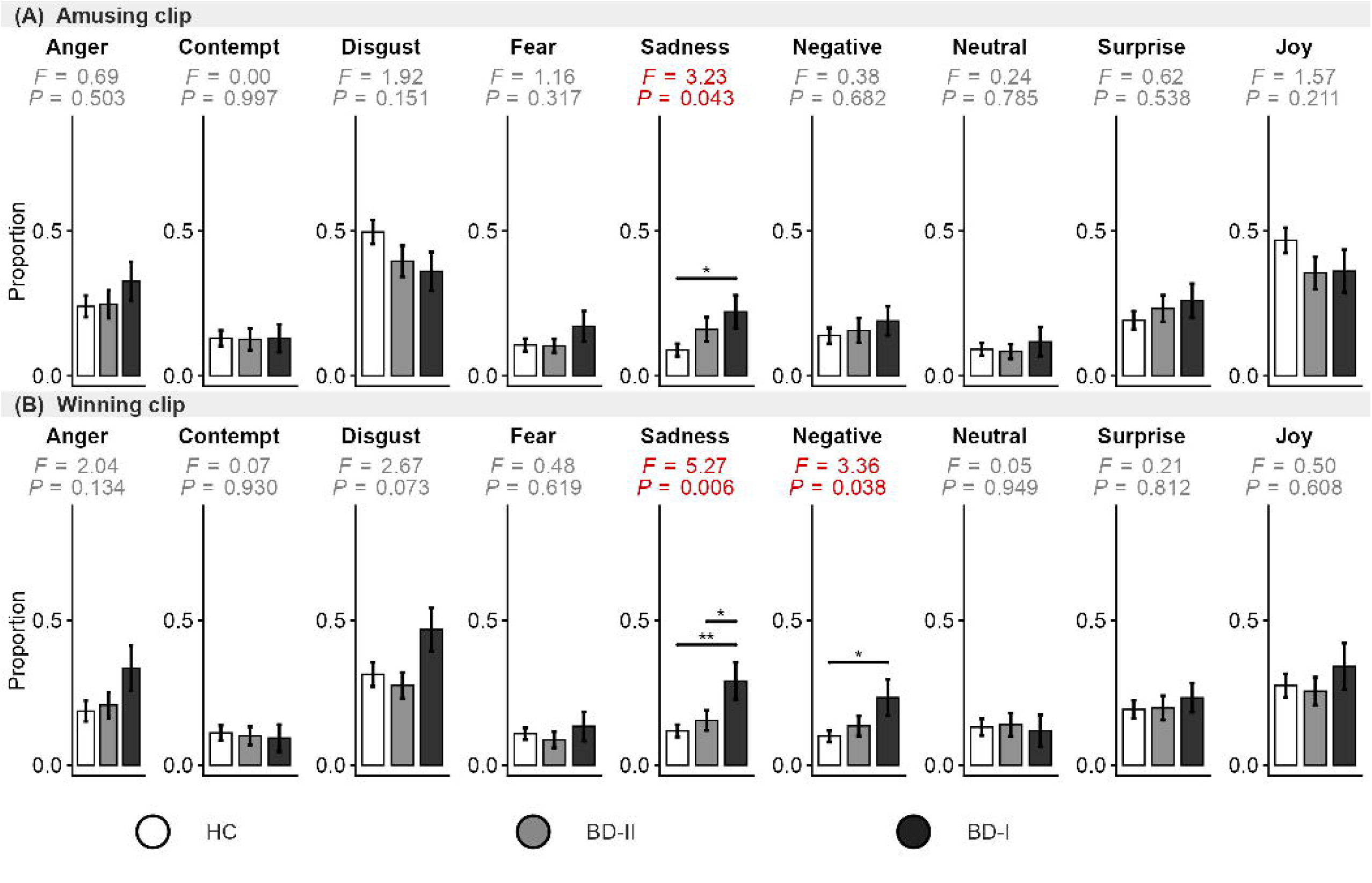
Facial expression proportions during amusing and winning film clips. Bar graphs show the mean values of the groups with standard error bars for each emotion channel during (A) an amusing film clip, and (B) a winning film clip. The dependent variable is the proportion of frames in which the baseline-corrected facial expression probability score equaled or exceeded 0.5. Significant between-group differences (one-way ANOVA, p < .05) are indicated by red F and p values; significant post-hoc pairwise comparisons (Tukey HSD) are denoted by brackets (* p < .05, ** p < .01, *** p < .001). BD-I, bipolar disorder type I; BD-II, bipolar disorder type II.

In addition to sadness, BD-I participants showed elevated anger-related responding in selected contexts. During the sad clip (**Figure 1A**), anger responses were significantly more frequent in BD-I than in HCs (F(2, 132) = 4.52, p = 0.013, η²p = 0.064; BD-I: M = 0.511, SD = 0.386; HCs: M = 0.258, SD = 0.330; p = 0.009), as were disgust responses (F(2, 132) = 4.68, p = 0.011, η²p = 0.066; BD-I: M = 0.556, SD = 0.329; HCs: M = 0.295, SD = 0.355; p = 0.008). In the neutral clip (**Figure 1B**), BD-I participants also displayed anger responses significantly more frequently than HCs (F(2, 132) = 4.91, p = 0.009, η²p = 0.069; BD-I: M = 0.400, SD = 0.368; Control: M = 0.164, SD = 0.283; p = 0.009). Across these comparisons, BD-II participants generally showed intermediate values and did not differ significantly from either BD-I or HCs.

A more differentiated pattern emerged in the socially anxious clip (**Figure 3A**), in which group differences were observed not only in negative emotion channels, but also in joy. In this context, BD-I participants showed significantly higher proportions of anger, fear, and joy responses than HCs. Group differences were significant for anger (F(2, 132) = 3.63, p = 0.029, η²p = 0.052; BD-I: M = 0.532, SD = 0.335; HCs: M = 0.315, SD = 0.316; p = 0.023), fear (F(2, 132) = 3.19, p = 0.044, η²p = 0.046; BD-I: M = 0.192, SD = 0.280; HCs: M = 0.087, SD = 0.145; p = 0.038), and joy (F(2, 132) = 3.35, p = 0.038, η²p = 0.048; BD-I: M = 0.398, SD = 0.412; HCs: M = 0.205, SD = 0.278; p = 0.043). BD-II did not differ significantly from HCs on any channel in this context. A related finding emerged in another threat-relevant context: in a clip depicting an anger scenario (**Figure 3B**), BD-I participants showed significantly more frequent fear responses than HCs (F(2, 132) = 4.03, p = 0.020, η²p = 0.057; BD-I: M = 0.150, SD = 0.240; HCs: M = 0.049, SD = 0.109; p = 0.015).

**Figure 3.**
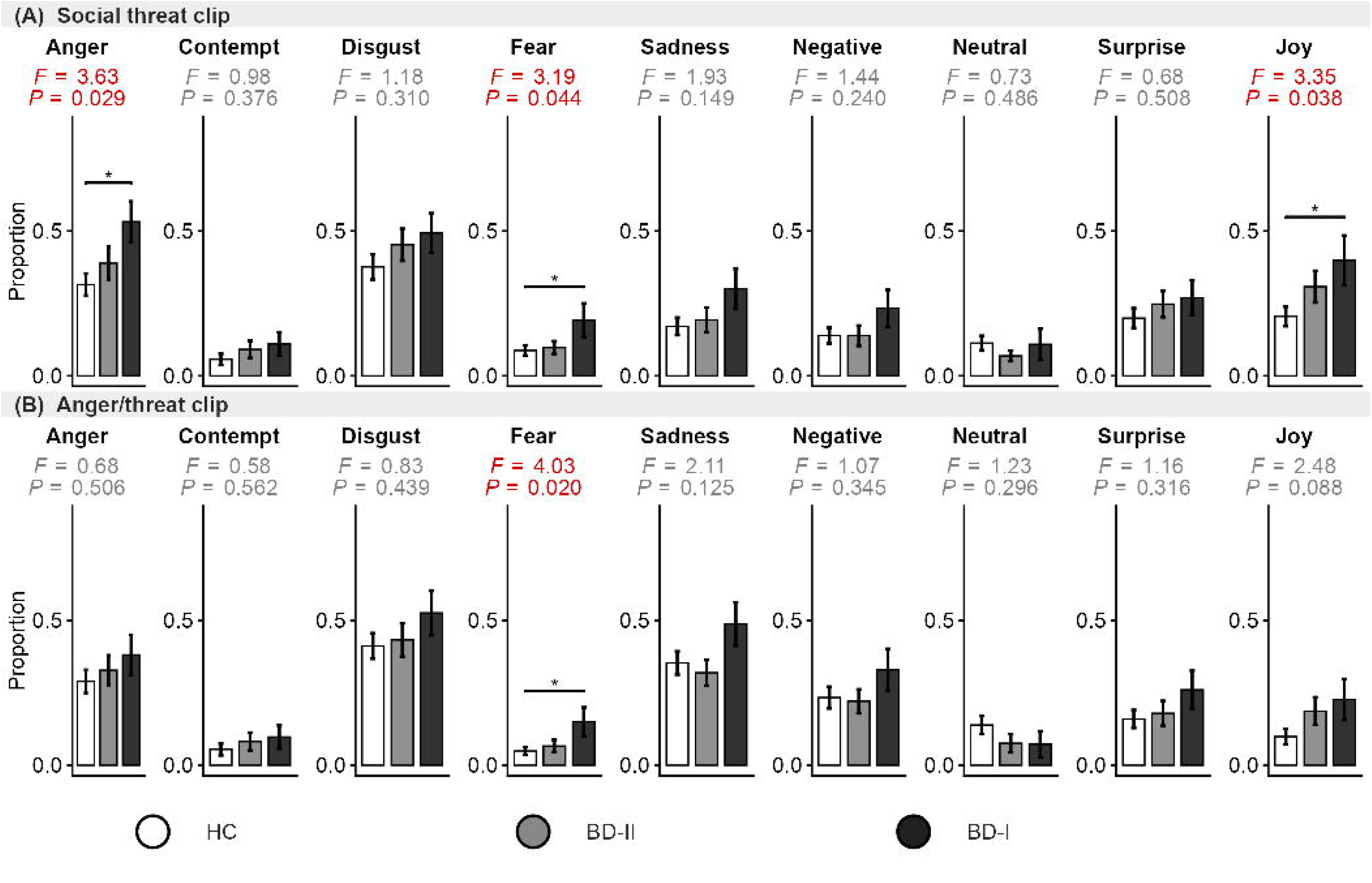
Facial expression proportions during social threat and anger film clips. Bar graphs show the mean values of the groups with standard error bars for each emotion channel during (A) a social threat film clip and (B) an anger/threat film clip. The dependent variable is the proportion of frames in which the baseline-corrected facial expression probability score equaled or exceeded 0.5. Significant between-group differences (one-way ANOVA, p < .05) are indicated by red F and p values; significant post-hoc pairwise comparisons (Tukey HSD) are denoted by brackets (* p < .05, ** p < .01, *** p < .001). BD-I, bipolar disorder type I; BD-II, bipolar disorder type II.

Finally, no significant group differences were observed during the racing, risk-taking, or happy clips (**Supplementary Figure 1**). Joy during the happy clip was numerically lower in both patient groups than in HCs (HCs: M = 0.594; BD-I: M = 0.472; BD-II: M = 0.477), although this difference did not reach significance (p = 0.218). Across the clip-level analysis, significant pairwise differences were observed only in contrasts involving BD-I; no significant BD-II versus HCs differences were found.

### 3.4. Supplementary analysis: linear mixed-effects model

To account for the repeated-measures structure of the clip-level data, supplementary linear mixed-effects analysis was conducted for each emotion channel. Significant Group × Video interactions were observed for joy (F(16, 1056) = 3.42, p < 0.001), disgust (F(16, 1056) = 3.35, p < 0.001), anger (F(16, 1056) = 2.15, p = 0.005), and sadness (F(16, 1056) = 1.81, p = 0.026). Follow-up pairwise comparisons were broadly consistent with the video-level ANOVA findings, with all significant post-hoc comparisons involving the BD-I group, whereas BD-II did not differ significantly from HCs in any video. Results were unchanged after the inclusion of demographic/clinical variables as covariates.

## 4. Discussion

The present study examined whether patients with BD would show distinguishable patterns of facial expression during emotional film clips using computer vision techniques. Relative to HCs, BD-I participants showed increased negative and context-incongruent facial responses across a range of affective film contexts. By contrast, BD-II participants did not differ significantly from HCs despite comparable depressive status. Importantly, the pattern observed in BD-I was not consistent with a simple account of generalized reduction in emotional responses, as abnormalities varied across emotional contexts, rather than appearing as a uniform reduction or increase in facial expressivity. Taken together, these findings suggest that facial-expression abnormalities during bipolar depression may differ across bipolar subtypes and reflect a distinct, context-sensitive disturbance in BD-I.

A robust finding was the amplified negative facial responding in BD-I across negative or threat-related contexts. In the sad clip, BDLI participants showed increased sadness together with elevated disgust and anger, suggesting a broader recruitment of negative affect, which extends prior findings of negatively biased facial expressions and selfLreports across diverse emotional film clips in patients with remitted BD (Kjærstad et al., 2020). Fear was also elevated in the threat-related clip, which may reflect heightened or generalized threat sensitivity in BD-I and is consistent with our related eye-tracking finding, that patients with BD-I showed greater selective attention to threatening stimuli than patients with BD-II (Sim et al., manuscript submitted for publication). Elevated anger during the neutral clip was likewise notable because it emerged in the absence of explicit emotional provocation. This pattern extends previous findings of emotional hyperreactivity to neutral stimuli in euthymic BD, (Dubois et al., 2012) and attentional biases toward threatening faces in symptomatic BD, including manic and depressive episodes (Gago et al., 2022), to patients with BD-I depression. Relatedly, youths with BD have been reported to perceive greater hostility and fear when viewing neutral faces (Rich et al., 2006). Considered together, these findings suggest that BD-I depression may involve not only intensified negative expression in clearly aversive situations, but also a tendency for negative emotional responding to emerge in ambiguous or minimally emotional contexts.

Patients with BD-I also showed facial responses that appeared poorly aligned with positive or rewarding contexts, most notably elevated sadness and a higher negative composite score during the winning clip compared to HCs. This pattern may reflect context-incongruent negative facial responding during reward-related stimulation, broadly in line with evidence of reward-processing abnormalities and difficulties sustaining positive affect in bipolar depression and related mood disorders (Long et al., 2022; Whitton et al., 2015; Whitton & Pizzagalli, 2022). In the present study, this disturbance was expressed not as a simple absence of emotional response, but as the intrusion of negative facial output into contexts that would ordinarily be expected to evoke more positive expression. Thus, the present finding should not be interpreted simply as reduced positive affect, but rather as context-incongruent negative facial responding during reward-related stimulation. In this respect, the current findings complement prior reports of reduced positive self-reported affect during winning and happy clips in patients with remitted BD (Kjærstad et al., 2020).

A qualitatively distinct multivalent pattern emerged in the socially anxious clip, in which patients with BD-I showed simultaneous elevations in anger, fear, and joy relative to HCs. Unlike the findings described above, which mainly reflected heightened negative responding in mood-congruent or context-inappropriate settings, this result was notable for the co-occurrence of negative and positive facial expressions within the same context. The elevation of joy in this clip is not readily explained by a simple reduction of emotional responses or anhedonia account, and instead suggests reduced contextual control or poor coordination of emotional expression. One possible interpretation is that socially anxious context may evoke not only heightened distress and anxiety-related responding, but also incongruous positive facial expression, perhaps reflecting reduced inhibition of context-inappropriate expression or unstable affective responding. This shares some similarities with prior work suggesting context-sensitive disturbances in emotional responding in BD across mood states rather than a uniform dampening of emotional output (Johnson et al., 2016; Townsend & Altshuler, 2012). At the same time, this finding should be interpreted cautiously. This interpretation should be regarded as tentative because the present measure does not resolve the temporal ordering or co-occurrence of these expressions.

In contrast to the pattern observed in BD-I, patients with BD-II did not differ significantly from HCs on any emotion channel at either the overall level, or the individual video level. This null finding suggests that facial-expression abnormalities during bipolar depression may not be uniform across subtypes and highlights the value of examining BD-I and BD-II separately rather than treating BD as a single diagnostic group (Brancati et al., 2023; Judd et al., 2003, 2005). Notably, the absence of significant effects in BD-II cannot be attributed solely to lower power, given that the BD-II group was larger than the BD-I group (n = 43 vs. n = 23). However, these findings should not be taken to indicate an absence of disturbance in emotional responding in BD-II depression. Rather, they may suggest that, under the current paradigm, such disturbance was less likely to manifest as overt facial output detectable by automated facial coding. Accordingly, the null finding in BD-II may be better understood not as an evidence of preserved emotional functioning, but as an indicator of subtype differences in bipolar depression, that may extend to how affective disturbance is expressed behaviorally. These findings raise the possibility that facial-expression patterns during naturalistic emotional film viewing may serve as candidate behavioral markers for differentiating BD-I and BD-II depression.

To the best of our knowledge, this is the first study to examine facial-expression abnormalities in patients with bipolar depression while explicitly distinguishing between BD-I and BD-II. By comparing these subtypes among patients assessed during a current depressive episode, the present design allowed to test whether facial-expression abnormalities represent a general feature of bipolar depression, or instead show subtype-specific patterns in the depressive episode. The use of automated facial-expression analysis during emotionally diverse film clips further enabled us to characterize these abnormalities across distinct affective contexts, supporting the ecological validity of this approach as a context-sensitive behavioral measure.

Several limitations should be considered. First, the present design cannot determine whether the observed differences between patients with bipolar depression and HCs reflect state-related effects of the depressive episode or more stable trait-like differences associated with bipolar disorder. Longitudinal follow-up across mood states will be needed to address this question. Second, mixed features were not directly assessed, limiting the study’s ability to determine whether the distinctive BD-I pattern was partly influenced by manic or hypomanic symptoms accompanying the depressive episode. Third, the facial-expression outcome was based on the proportion of threshold-exceeding frames for each emotion, which did not allow to determine in detail, whether different expressions co-occurred or alternated over time. Fourth, the sample consisted of Korean participants and the film clips were selected from Korean-language materials. The generalizability of the present findings to other cultural groups should be tested in future studies because context-appropriate emotional responses and expression may vary across cultures. Thus, the present findings should be regarded as an initial characterization of subtype-related differences in facial responses during bipolar depression.

In conclusion, the present findings suggest that facial-expression abnormalities during bipolar depression may vary by both emotional context and bipolar subtype. Rather than showing a uniform reduction or increase in facial expressivity, patients with BD-I exhibited a pattern characterized by heightened negative reactivity, reduced context-appropriate modulation of emotional expression, and, in some contexts, potentially incongruous affective responding. By contrast, patients with BD-II did not show comparable abnormalities on the present facial-expression measures, highlighting the importance of distinguishing bipolar subtypes rather than treating BD as a single diagnostic group. These findings support the value of context-sensitive behavioral assessment using naturalistic emotional stimuli and suggest that automated facial-expression analysis may provide a useful adjunctive measure for characterizing affective disturbance in bipolar depression. Future longitudinal studies that directly assess mixed features across mood states will be important for clarifying the clinical significance and diagnostic specificity of these findings.

## Data Availability

All data produced in the present study are available upon reasonable request to the authors

## Acknowledgements

We are grateful to Eunhye Jeong, M.A., and Young Hyun Cho, B.N., for contributions in conducting the experiments.

## Ethical standards

The authors assert that all procedures contributing to this work comply with the ethical standards of the relevant national and institutional committees on human experimentation and with the Helsinki Declaration of 1975, as revised in 2008.

---

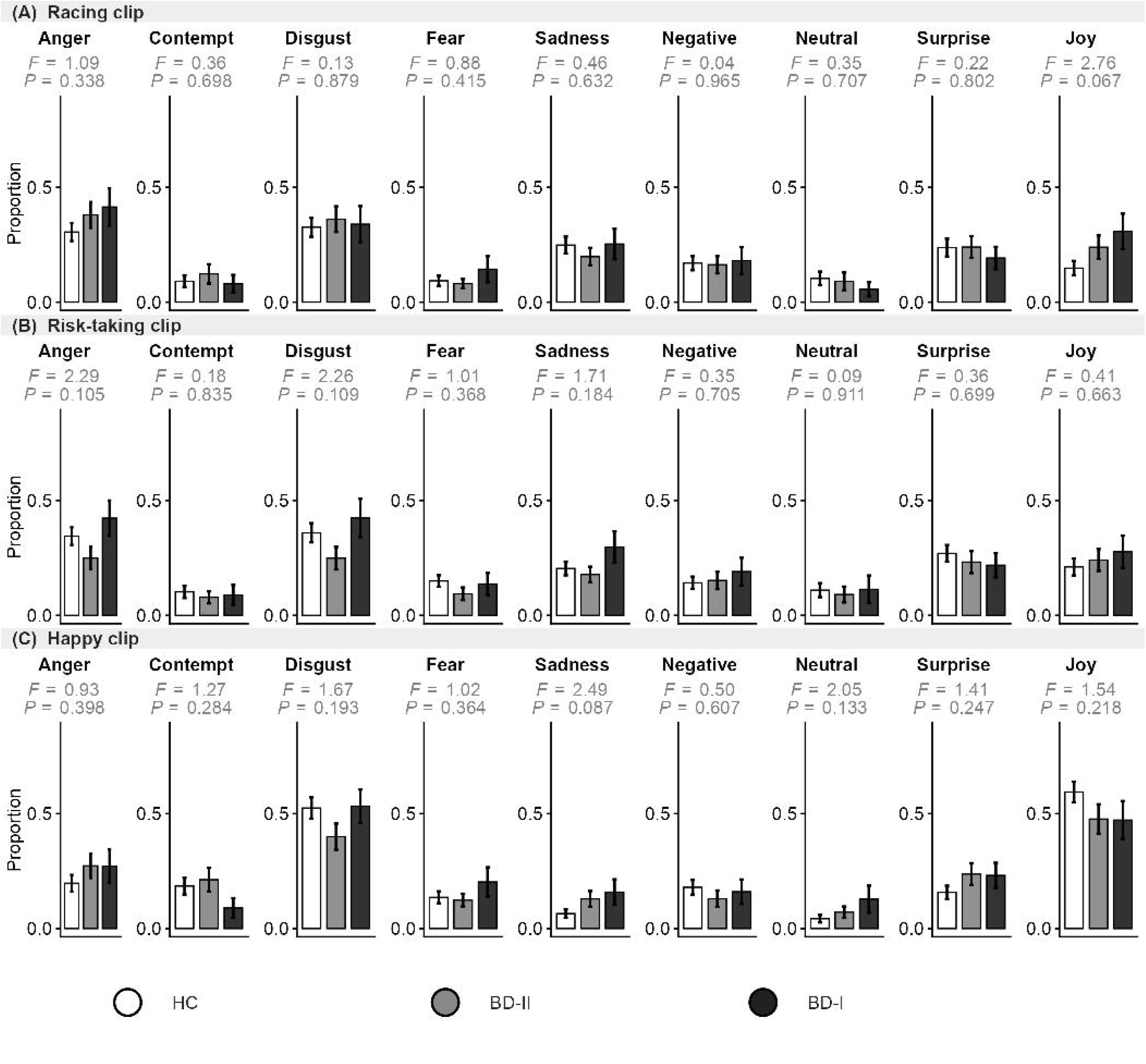

