## Supplementary Materials for "Context-dependent facial-expression patterns during affective film viewing in patients with bipolar depression"

**Supplementary Material 1**. Measures

Patient Health Questionnaire-9 (PHQ-9)

Depressive symptom severity was measured using the Patient Health Questionnaire-9 (PHQ-9), a nine-item self-report scale developed to assess depressive symptoms over the preceding two weeks (Kroenke et al., 2001; Shin et al., 2010). Each item is rated from 0 (not at all) to 3 (nearly every day), yielding a total score between 0 and 27, with higher scores reflecting more severe depressive symptoms. In the present sample, internal consistency was acceptable, with Cronbach’s α values of .768 in the BD group and .756 in the HC group.

Generalized Anxiety Disorder-7 (GAD-7) Scale

Anxiety symptom severity was assessed using the Generalized Anxiety Disorder-7 scale (GAD-7), a seven-item self-report measure of anxiety symptoms experienced during the past two weeks (Seo et al., 2014; Spitzer et al., 2006). Items are scored on a four-point scale ranging from 0 (not at all) to 3 (nearly every day), with higher total scores indicating greater anxiety severity. Cronbach’s α values in the current study were .845 for the BD group and .804 for the HC group, indicating good internal consistency.

Columbia-Suicide Severity Rating Scale (C-SSRS)

Suicidal ideation was assessed using the Columbia-Suicide Severity Rating Scale (C-SSRS), a clinician-administered semi-structured interview designed to evaluate suicidal ideation and behavior (Jang et al., 2014; Posner et al., 2011). The present study used the severity subscale for suicidal ideation, which consists of five ordered items assessing increasing severity of suicidal thoughts. When a participant endorsed one or more items, the highest endorsed item was used as the severity score, with higher scores indicating more severe suicidal ideation.

**Supplementary Material 2.** Stimulus-selection procedure for the emotional film clips

1. Overview

To develop a set of emotional film stimuli suitable for Korean participants, a separate validation procedure was conducted before the main experiment. The aim was to identify Korean-language clips that would reliably evoke the intended emotional responses while maintaining the general logic of the film-clip paradigm used in prior work. Therefore, candidate clips were screened and empirically evaluated before inclusion in the final stimulus set.

2. Candidate clip selection

Candidate clips were identified from Korean films, television programs, or other audiovisual materials suitable for Korean participants. For most target categories—amusement, anger, happiness, neutrality, sadness, racing, risk-taking, and winning—two candidate clips were initially selected. For the socially anxious category, only one candidate clip was included because a suitable alternative clip meeting the selection criteria could not be identified. Candidate clips were chosen to depict the intended emotional context clearly, to be understandable without extensive background information, and to avoid excessively violent, sexually explicit, or otherwise inappropriate content.

3. Independent validation sample

An independent validation sample of 31 Korean-speaking adults participated in the clip-evaluation procedure. The sample included 14 men and 17 women, with a mean age of 30.03 years. Participants with a history of psychiatric diagnosis were not included in the validation sample. None of the participants in this validation procedure took part in the main experiment.

4. Rating procedure

Candidate clips were presented in a randomized order. For clips targeting amusement, anger, happiness, neutrality, and sadness, participants first indicated the emotion they experienced most strongly while watching the clip by selecting one response from the following options: happiness, sadness, anger, surprise, disgust, fear, amusement, or neutrality/no clear emotion. This was followed by rating the overall intensity of the emotion they experienced on a seven-point scale ranging from 1 (very weak) to 7 (very intense). For the racing, risk-taking, and winning clips, the forced-choice emotion-identification item was omitted; participants only rated the overall intensity of their emotional response using the same seven-point scale.

5. Selection criteria

For clips targeting amusement, anger, happiness, neutrality, and sadness, the final stimulus was selected using two criteria: the proportion of participants who identified the intended emotion as the dominant emotional response, and the mean intensity rating for the emotion experienced while watching the clip. When these two criteria favored different candidate clips, priority was given to the higher intensity rating. For clips targeting racing, risk-taking, socially anxious, and winning contexts, selection was based solely on the mean intensity rating.

6. Validation results

**Supplementary Table 1** summarizes the validation results used to determine the final stimulus set.

7. Final stimulus set

The final selected clips and their brief narratives are reported in **Table 1** of the main text.

**Supplementary Table 1.** Validation results for candidate emotional film clips

| **Target category** | **Candidate** | **Intended-emotion agreement (%)** | **Mean intensity rating** | **Selected** |
| --- | --- | --- | --- | --- |
| Amusing | A | 71.0 | 3.50 | No |
|  | B | 80.7 | 3.61 | Yes |
| Anger | A | 64.5 | 5.45 | Yes |
|  | B | 25.8 | 4.65 | No |
| Happy | A | 80.7 | 3.97 | No |
|  | B | 80.7 | 3.97 | Yes |
| Neutral | A | 93.6 | 2.19 | Yes |
|  | B | 80.7 | 2.03 | No |
| Sad | A | 77.4 | 4.61 | Yes |
|  | B | 80.7 | 4.22 | No |
| Racing | A | — | 4.16 | Yes |
|  | B | — | 3.9 | No |
| Risk taking | A | — | 4.45 | Yes |
|  | B | — | 4.19 | No |
| Socially anxious | — | — | 4.55 | Yes |
| Winning | A | — | 4.51 | No |
|  | B | — | 4.52 | Yes |

*Note*. Agreement refers to the percentage of participants selecting the intended emotion as the dominant response. Intensity ratings ranged from 1 (very weak) to 7 (very intense). The forced-choice emotion-identification item was administered only for amusement, anger, happiness, neutrality, and sadness clips.
